# Severus: accurate detection and characterization of somatic structural variation in tumor genomes using long reads

**DOI:** 10.1101/2024.03.22.24304756

**Authors:** Ayse Keskus, Asher Bryant, Tanveer Ahmad, Byunggil Yoo, Sergey Aganezov, Anton Goretsky, Ataberk Donmez, Lisa A. Lansdon, Isabel Rodriguez, Jimin Park, Yuelin Liu, Xiwen Cui, Joshua Gardner, Brandy McNulty, Samuel Sacco, Jyoti Shetty, Yongmei Zhao, Bao Tran, Giuseppe Narzisi, Adrienne Helland, Daniel E. Cook, Pi-Chuan Chang, Alexey Kolesnikov, Andrew Carroll, Erin K. Molloy, Irina Pushel, Erin Guest, Tomi Pastinen, Kishwar Shafin, Karen H. Miga, Salem Malikic, Chi-Ping Day, Nicolas Robine, Cenk Sahinalp, Michael Dean, Midhat S. Farooqi, Benedict Paten, Mikhail Kolmogorov

**Affiliations:** Center for Cancer Research, National Cancer Institute, NIH, Bethesda, MD, USA; Children’s Mercy Hospital, University of Missouri-Kansas City School of Medicine, Kansas City, MO, USA; Oxford Nanopore Technologies, NY, USA; Department of Computer Science, University of Maryland, College Park, MD, USA; Division of Cancer Epidemiology and Genetics, National Cancer Institute, NIH, Rockville, MD, USA; UC Santa Cruz Genomics Institute, Santa Cruz, CA, USA; Sequencing Facility, Cancer Research Technology Program, Frederick National Laboratory for Cancer Research, Frederick, MD, USA; Sequencing Facility Bioinformatics Group, Biomedical Informatics and Data Science Directorate, Frederick National Laboratory for Cancer Research, Frederick, MD, USA; New York Genome Center, NY, USA; Google Inc, Mountain View, CA, USA

## Abstract

Most current studies rely on short-read sequencing to detect somatic structural variation (SV) in cancer genomes. Long-read sequencing offers the advantage of better mappability and long-range phasing, which results in substantial improvements in germline SV detection. However, current long-read SV detection methods do not generalize well to the analysis of somatic SVs in tumor genomes with complex rearrangements, heterogeneity, and aneuploidy. Here, we present Severus: a method for the accurate detection of different types of somatic SVs using a phased breakpoint graph approach. To benchmark various short- and long-read SV detection methods, we sequenced five tumor/normal cell line pairs with Illumina, Nanopore, and PacBio sequencing platforms; on this benchmark Severus showed the highest F1 scores (harmonic mean of the precision and recall) as compared to long-read and short-read methods. We then applied Severus to three clinical cases of pediatric cancer, demonstrating concordance with known genetic findings as well as revealing clinically relevant cryptic rearrangements missed by standard genomic panels.

## Introduction

Somatic structural variation (SV) is a hallmark of cancer, typically defined as a process that can insert, delete, or rearrange genomic sequences longer than 50 bp (Cosenza, Rodriguez-Martin, and Korbel 2022). In comparison to small variants (SNVs and short indels), somatic SVs vary greatly in size and complexity, from simpler deletions and focal amplifications to catastrophic events shuffling large fragments from one or multiple chromosomes (Stephens et al. 2011). Different cancer types often have characteristic patterns of somatic SVs (Y. Li et al. 2020) caused by various mutational processes (Carvalho and Lupski 2016) and overall genomic instability (Drews et al. 2022). The SV process plays a critical role in carcinogenesis; a recent analysis of 2,658 tumor genomes showed that ∼50% of driver mutations overlap with SVs (“Pan-Cancer Analysis of Whole Genomes” 2020).

Most current large-scale whole-genome sequencing projects rely on short-read sequencing (e.g., Illumina) to call germline and somatic SVs (Chen et al. 2015; Cameron et al. 2017; Wala et al. 2018; Rausch et al. 2012; Fan et al. 2014). In germline SV analysis, short-read methods often miss variants inside repetitive regions (such as variable number tandem repeats - VNTRs - or segmental duplications). As approximately half of the germline SVs coincide with repeats (Chaisson et al. 2019), recent benchmarks reported 30-70% recall rates for short-read methods (Zook et al. 2020; Wagner et al. 2022; Zarate et al. 2020).

Compared to germline SVs, somatic SVs in cancer may be driven by different mutational processes not involving repetitive DNA (Carvalho and Lupski 2016). As a result, a higher proportion of the somatic SV landscape is accessible to short reads, as compared to germline SVs (Choo et al. 2023). However, as we illustrate below, short-read methods still miss many somatic variants, in particular clustered SVs, insertions, and short SVs. We also demonstrate that short-read methods may have biased patterns of false-positive errors, which may be problematic for association studies.

Recent developments in long-read sequencing technologies, such as those produced by Oxford Nanopore Technologies (ONT) or Pacific Biosciences (PacBio), can address the limitations of the short-read SV calling methods (Jiang et al. 2020; Smolka et al. 2024; Sedlazeck et al. 2018). This is because of the better ability to disambiguate repeats (Logsdon, Vollger, and Eichler 2020) and to phase variants into megabase-scale haplotypes (Lin et al. 2022; Mahmoud et al. 2021; Shafin et al. 2021).

However, most of the existing long-read developments were focused on the analysis of germline variation rather than cancer genomes. Long-read sequencing has been recently applied to various cancers, including lung (Sakamoto et al. 2020, 2022), liver (Fujimoto et al. 2021), medulloblastoma (Rausch et al. 2023), and cervical (Rossi et al. 2023; Zhou et al. 2022; Akagi et al. 2023). Further, another recent study applied nanopore sequencing to 189 patient tumors across multiple cancer types (O’Neill et al. 2024). In the absence of specialized methods for somatic SV calling, the above studies utilized the existing germline methods; below, we illustrate that this may lead to suboptimal results.

In this work, we present Severus, a tool for somatic SV calling using long reads. Unlike the existing germline SV callers, Severus was optimized for complex SV patterns and abnormal karyotypes and supports input of matching normal samples and multiple tumor samples. Severus uses long reads to phase germline and somatic variants into haplotypes. Furthermore, Severus constructs a phased breakpoint graph to cluster SVs to complex rearrangement events (Hadi et al. 2020; Choo et al. 2023; Aganezov and Raphael 2020; Shale et al. 2022). These capabilities of phasing and characterizing complex rearrangements distinguish Severus from two other recently developed methods for long-read somatic SV calling: nanomonsv (Shiraishi et al. 2023) and SAVANA (https://github.com/cortes-ciriano-lab/savana).

Currently, only a limited number of datasets with long-read tumor sequencing data are available. To address that, we sequenced five tumor/normal cell line pairs with Illumina, ONT, and PacBio sequencing instruments and generated a set of confident somatic SV calls for each tumor cell line. These data and benchmarks are being made openly available to encourage future method development.

Using these cell line benchmarks, we show that Severus outperforms existing long-read and short-read methods in somatic SV detection across the different datasets and cancer types. We also sequence three clinical cases of pediatric cancer and use Severus to identify driver events and clinical genetic subtypes.

## Results

### Overview of the Severus algorithm

To detect and characterize SVs, we use a popular model in which an SV is represented by one or multiple two-breakpoint junctions. Each junction connects two non-adjacent reference positions called breakpoints (also commonly referred to as breakends). Each breakpoint is defined by its chromosomal coordinate and direction (5’ or 3’). We denote simple SVs as the ones commonly observed in the germline: insertions, deletions, duplications, and reciprocal inversions (Figure 1). Various mutational processes in cancer may result in more complex SVs defined by multiple junctions (e.g., chromoplexy, chromothripsis, templated insertions, and others; Figure 1). The goal of Severus is (i) to accurately identify and classify individual junctions and (ii) to cluster junctions into complex SVs.

**Figure 1.**
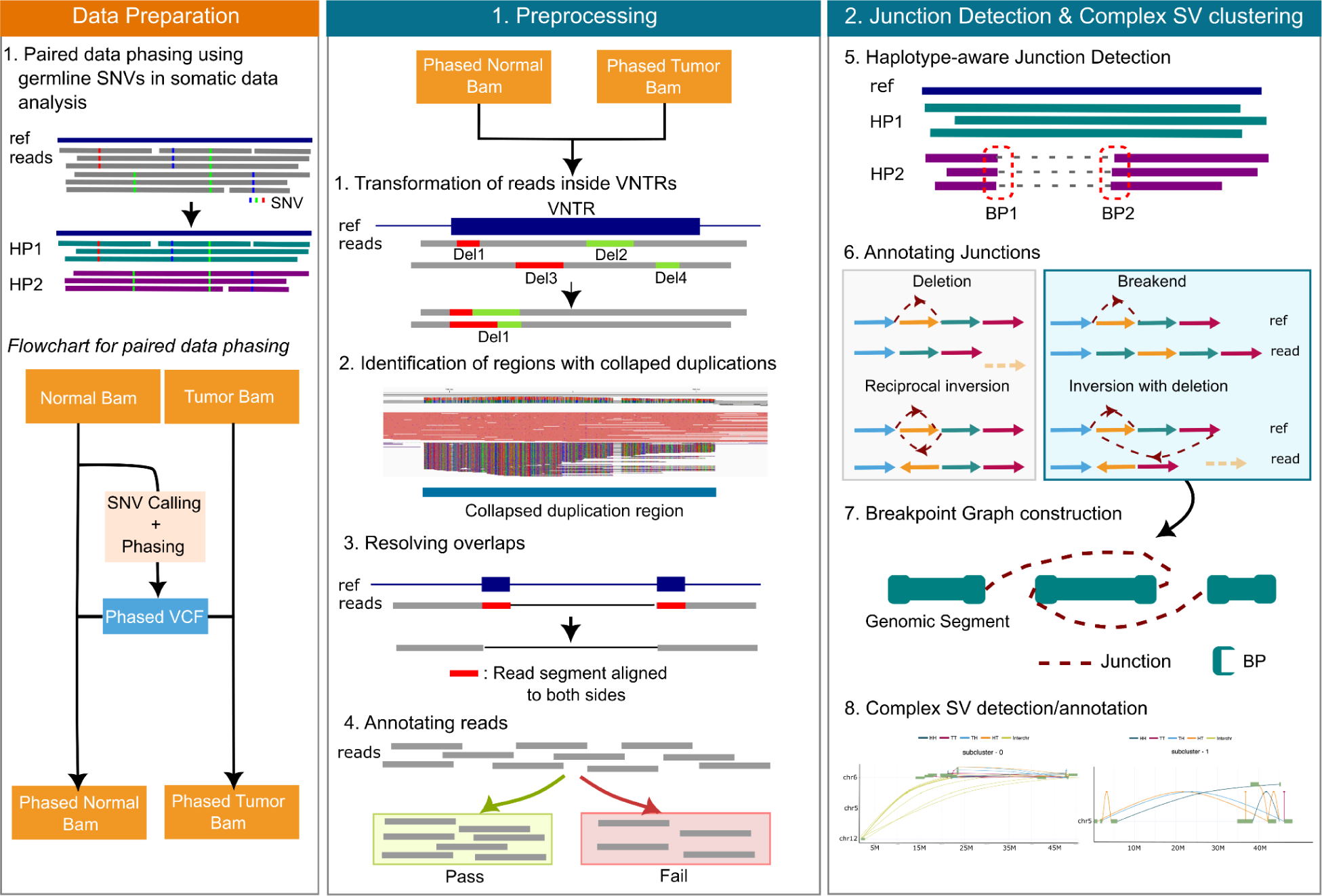
An overview of the Severus algorithm. (A) Phasing starts with SNV calling and phasing using aligned normal data; phased VCF is then used to haplotag tumor and normal alignments. (B) Severus has multiple pre-processing steps: (i) handling unstable alignment of reads with indels in VNTR regions, allowing uniform representation of the indels, (ii) identification of misaligned reads to collapsed duplication regions, (iii) resolving overlapping alignments of same read segments on both sides of the junctions (iv) annotating reads as pass or fail. (C) Severus calls haplotype-aware junctions from split alignments, then identifies simple junctions (indels and reciprocal inversions). Severus constructs a breakpoint graph with the rest of the junctions and identifies complex SVs.

To call junctions, Severus analyzes input long-read alignment from the tumor and normal samples. The pipeline first calls germline SNPs in a normal sample, phases SNPs, and assigns a haplotype to each aligned read in both normal and tumor samples. Severus then processes haplotyped alignments by identifying collapsed repeat regions and unifying alignment coordinates inside the tandem repeat regions. Severus jointly analyzes all input samples to generate an initial set of SVs and then genotypes each variant inside individual samples.

To cluster junctions and classify SVs, Severus builds a phased breakpoint graph (Figure 1). The graph preserves phasing information and implicitly encodes the derived structure of tumor haplotypes. Complex SVs are then detected and annotated as subgraphs of the breakpoint graph. We note the similarities of our approach with other graph-based methods for merging SV calls into haplotypes, such as Jabba (Hadi et al. 2020; Choo et al. 2023), RCK (Aganezov and Raphael 2020), LINX (Shale et al. 2022), InfoGenomeR (Lee and Lee 2021) and others. In contrast to these methods, Severus (i) implements additional constraints based on long-read phasing and (ii) simplifies the graph using long-read connectivity.

### Evaluating SV calling performance using Minda

We first sought to benchmark Severus using various cell line sequencing datasets and compare it against the other popular short-read and long-read approaches. We selected three popular Illumina-based somatic SV callers, Manta (Chen et al. 2015), SVaBA (Wala et al. 2018), and GRIDSS/GRIPSS (Cameron et al. 2017), as well as two recently developed long-read somatic SV callers, nanomonsv (Shiraishi et al. 2023) and SAVANA (https://github.com/cortes-ciriano-lab/savana). We also evaluated Sniffles2 (Smolka et al. 2024), a popular long-read germline SV caller. Although Sniffles2 was not designed for a paired tumor-normal analysis, we used its mosaic mode to subtract the normal sample SV calls from the tumor sample calls.

Benchmarking of SVs is challenging because different tools may represent the same SVs differently. For example, a tandem duplication may be represented as an insertion or as a breakend (BND; junction) call. Current SV evaluation methods, such as truvari (English et al. 2022), were primarily designed for the comparison of indels, the most common type of germline SVs. We, therefore, developed a new comparison tool called Minda, which is agnostic to SV types and is better suited for analysis of somatic SVs represented as junctions. Minda can also be used for multiway call sets comparison and merging (Jeffares et al. 2017; Kirsche et al. 2023).

### Benchmarking long-read tools using the GIAB HG002 germline SV set

Complete and accurate detection of germline SVs is a prerequisite for somatic SV calling. Thus, we first benchmarked Severus along with Sniffles2 (Smolka et al. 2024) and cuteSV (Jiang et al. 2020) against the recent germline SV benchmark for the HG002 genome (Zook et al. 2020) with ∼40x ONT R10 sequencing data as input (Kolmogorov et al. 2023) (Figure 2, Supplementary Table 1). Reassuringly, Severus had the highest F1-score (0.9698) along with Sniffles2 (0.9663), followed by cuteSV (0.9422). As SAVANA and nanomonsv require matching normal data as input, we did not evaluate them in this benchmark.

**Figure 2.**
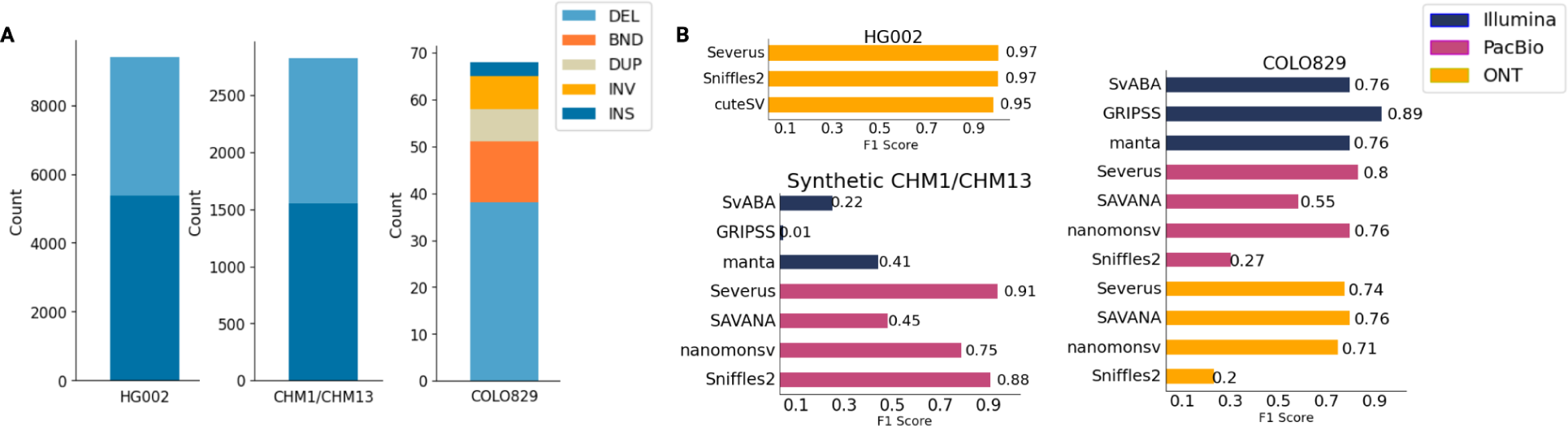
Benchmarking of Severus and other SV callers with the existing benchmarking sets. (a) SV numbers and types for each benchmark and (b) the corresponding F1 scores. HG002 represents the GIAB HG002 SV benchmark; CHM1/CHM13 represents a synthetic mix of CHM1 and CHM13 PacBio HiFi data, with CHM1 sequencing as normal. Both HG002 and CHM1/CHM13 are computed inside the GIAB HG002 Tier1 confident regions. The COLO829 benchmark is evaluated against the Valle-Inclan et al. (2022) call set. F1 scores are computed using Minda. DEL = Deletion, BND = breakend junction, DUP = duplication, INV = inversion, INS = insertion.

### Benchmarking using a synthetic mixture of the CHM1 and CHM13 genomes

Next, we benchmarked short- and long-read tools using a mock tumor-normal dataset with known ground truth. Similar to the earlier study (H. Li et al. 2018), we utilized two hydatidiform mole cell lines (CHM1 and CHM13) that are effectively haploid and lack allelic heterozygosity. We combined PacBio HiFi sequencing data of CHM1 and CHM13 in equal proportions to mimic a tumor sample; sequence data from only CHM1 represented the normal sample. The correct set of “somatic” SV calls were SVs that were unique to CHM13 and did not appear in CHM1. The ground truth set of SV calls was computed from the CHM1 and CHM13 assemblies (Nurk et al. 2022; Steinberg et al. 2014) using hapdiff (Kolmogorov et al. 2023).

In the benchmark, Severus and Sniffles2 performed the best (F1 scores 0.9145 and 0.8758, respectively), followed by nanomonsv (0.7456). SAVANA had a lower F1 score (0.4349), primarily explained by its reduced precision (Figure 2, Supplementary Table 1). Consistent with the germline SV benchmarks, Illumina-based tools showed low F1 scores (ranging from 0.01 to 0.4). Since the CHM1/CHM13 benchmark is based on germline SVs, it predominantly consists of indels, approximately half of which are inside VNTR regions (Chaisson et al. 2019); these are known limitations of Illumina-based short-read methods.

### Benchmarking using the curated set of somatic SVs in the COLO829 melanoma cell line

A recent study has constructed and validated a confident set of 68 somatic SV calls for the COLO829 melanoma cell line (Espejo Valle-Inclan et al. 2022). To evaluate short- and long-read methods using this benchmarking call set, we sequenced the COLO829/COLO829BL cell lines using Illumina and ONT R9 protocols at 100x for both tumor and normal samples (Supplementary Table 2). In addition, we downloaded publicly available PacBio HiFi data (https://www.pacb.com/connect/datasets/).

Previous studies have shown that the COLO829 cell line is heterogeneous and can contain distinct clones (Velazquez-Villarreal et al. 2020). As a result, false-positive calls against the benchmarking set may represent rare/emerging subclonal variants. To account for that, we only considered calls with at least a 10% variant allele frequency (VAF) for all tools (Figure 2; Supplementary Table 3).

On the PacBio dataset, Severus had the highest F1 score (0.79), followed by nanomonsv (0.76) and SAVANA (0.53). On the ONT R9 dataset, SAVANA had the highest score (0.75), followed by Severus (0.74) and nanomonsv (0.69). Sniffles2 had reduced F1 scores on both PacBio and ONT datasets (0.262 and 0.1967, respectively), which was in contrast to the substantially higher F1 scores of Sniffles2 on the CHM1/CHM13 benchmark. This highlights the benefit of a specialized approach for somatic SV calling. Illumina methods performed substantially better on COLO829 (F1-scores ranging from 0.76 to 0.89), as compared to the CHM1/CHM13 benchmark; this is likely explained by the much lower proportion of somatic insertions.

GRIPSS F1-score (0.89) was higher than the scores of all long-read methods; primarily due to nearly-perfect precision. It is possible that the current version of the benchmark may miss some real subclonal variants (called by other tools but not GRIPSS). We manually examined false positive calls made by Severus, and 60% of them were subclonal SVs with unambiguous read support. These insights are in line with the recent re-evaluation of the COLO829 benchmark (Paulin et al. 2024).

### Establishing a multi-platform benchmark using a tumor/normal cell line panel

A limitation of the COLO829 benchmark is the relatively small number of somatic SV calls. With the goal of creating an additional benchmarking resource, we generated a multi-omics panel of five additional tumor cell lines and their corresponding matched normals: HCC1954, H2009, HCC1937, H1437, and HCC1395. The first four cell line pairs were sequenced from the same DNA extract using Illumina, ONT R10, and PacBio HiFi with high coverage (Figure 3A, Supplementary Table 2). For HCC1395, we only performed ONT R10 sequencing, and existing Pacbio HiFi and Illumina data was downloaded from the public repositories (Fang et al. 2021) (https://www.pacb.com/connect/datasets/). The panel represents two lung and four breast cancer cell lines, with the number of somatic SVs varying from 200 to 1300 (Figure 3B).

**Figure 3.**
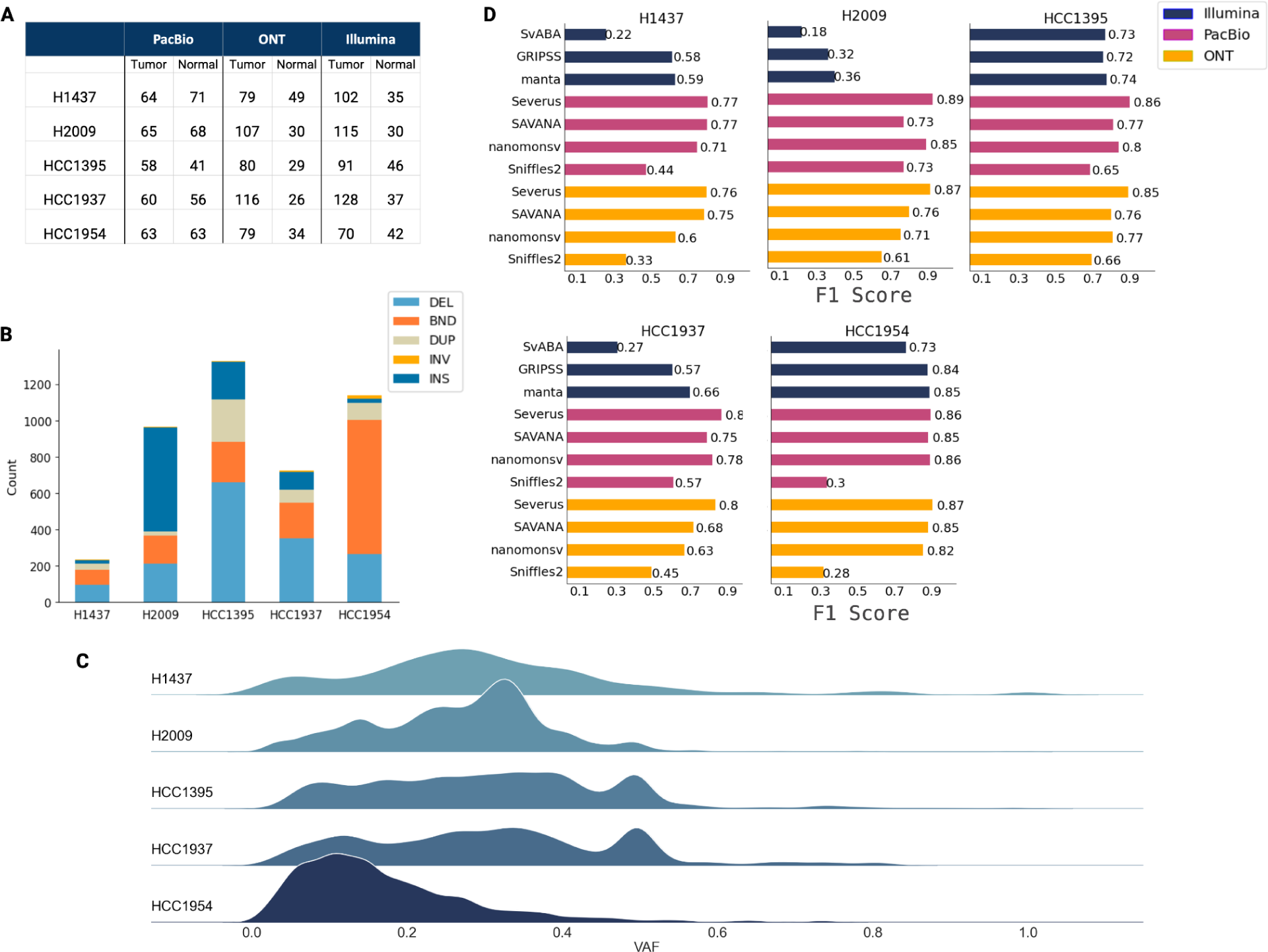
Benchmarking Severus and other SV callers using a multi-technology cell line panel. Confident set SVs are supported by at least 2 out of 3 technologies and 4 out of 11 call sets. F1 scores are evaluated against the confident sets using Minda. (A) Sequencing depth for each cell line and technology. (B) Number of confident SVs and their types. (C) Variant allele fraction of confident SVs. (D) F1 scores of each tool, grouped by sequencing technology.

In contrast to the COLO829 benchmark, there were no existing SV benchmarks for most of the newly sequenced cell lines; a previous study has produced a benchmarking SV set for the HCC1395 cell line but with earlier versions of PacBio and ONT protocols (Talsania et al. 2022). We thus created an ensemble of confident calls by merging the results from different methods and sequencing technologies using Minda. We define a call as confident if it is supported by at least two (out of three) technologies and at least four (out of 11) callers. SV types for the SVs in the ensemble call are defined with majority voting. Minda also outputs the median VAF calculated for each tool using the number of support and spanning reads.

To validate the ensemble benchmarking strategy, we constructed an ensemble call set for the COLO829 dataset. The corresponding F1 scores for each tool were highly correlated with the F1 scores against the Valle-Inclan et al. benchmark (Spearman’s r = 0.90; Supplementary Figure 1, Supplementary Table 4).

For datasets other than HCC1395, Illumina sequencing initially had substantially higher depth compared to the long-read data. This resulted in an increased number of subclonal SV calls, unsupported by other technologies and marked as false positives. We thus downsampled Illumina tumor sequencing to approximately 100x depth. Because sequencing for all cell lines (except HCC1395) was derived from the same DNA extraction, clonal distribution is expected to be consistent; we thus did not filter variants with low VAF in this benchmark (variants with low VAF are typically harder to detect, as we illustrate below).

The SV ensemble parameters were selected to balance between precision and recall, and we did not observe the resulting ensemble calls to be overly sensitive to the Minda parameters (Supplementary Table 5).

Each cell line had a unique combination of different SV types. H1437 had the least number of somatic SVs. In the two cell lines with the highest number of somatic SVs, HCC1395 consisted mainly of indels, whereas HCC1954 - in which the consistency between technologies was the highest - had an elevated number of junctions (Supplementary Figures 2, 3). In the other lung cancer line, H2009, insertions were the most abundant, and it is the only cell line that the majority of the calls (60%) were supported by only long-read sequencing (Supplementary Figures 2, 3). For most cell lines, the VAF of SV calls was evenly distributed between 0 and 0.5, suggesting the presence of subclonal variants and aneuploidy (Figure 3C). As an exception, HCC1954 had an increased number of SVs with VAF < 0.25, a signature of genome doubling.

### Benchmarking algorithms and technologies against the ensemble truth set

On this benchmark, Severus consistently produced the highest F1 scores (ranging from 0.76 to 0.89) in the PacBio and ONT categories across all cell lines (Figure 3D, Supplementary Table 6). On most cell lines, SAVANA was the second-best in the ONT category, and nanomonsv was second-best in the PacBio category. Sniffles2 had consistently reduced F1 scores, yet it performed better in cell lines HCC1395 and H2009, with a higher prevalence of indels.

In the Illumina category, Manta consistently had better F1 scores compared to GRIDSS and SVaBA (Figure 3D, Supplementary Table 6). Aggregating by technology, the Illumina methods had lower F1 scores compared to the PacBio and ONT methods on most datasets. HCC1954 was a notable exception: Manta and GRIPSS slightly outperformed SAVANA and nanomonsv on ONT data. Overall, although the typical VAF of SVs in HCC1954 was lower, the agreement between tools and technologies was the highest among the cell lines. On the other hand, Illumina tools had low F1 scores on the H2009 dataset with a high proportion of insertions.

Across cell lines, Severus SV calls for ONT and PacBio inputs were overall highly consistent with similar F1 scores. On most datasets, ONT-based calls had higher recall, while PacBio-based calls had higher precision (Supplementary Table 6). To evaluate if this is a consequence of different sequencing depths of tumor and normal samples for ONT and PacBio, we downsampled both technologies to 60x tumor / 30x normal coverage. We then evaluated the Severus calls on downsampled data against the ensemble truth set for the HCC1954 and H2009 cell lines (Supplementary Table 7). On both datasets, this resulted in even higher consistency between F1 scores (0.8673 for ONT, 0.8536 for PacBio on HCC1954; 0.8760 for ONT, and 0.8793 for PacBio on H2009). However, the tradeoff between precision and recall remained, albeit at a lower magnitude.

### Stratification of error patterns of different sequencing technologies and algorithms

The performance of all tools was variable on different datasets, which was primarily explained by the differences in SV patterns and genomic context around the SV breakpoints. We annotated each SV call with the potential challenges for the algorithms (Figure 4; Supplementary Table 8). The challenging categories can be loosely separated into (i) types of SV, e.g., insertion, (ii) local mappability and repeat context, (iii) SV size and allelic fraction, and (iv) SV clusters and chains.

**Figure 4.**
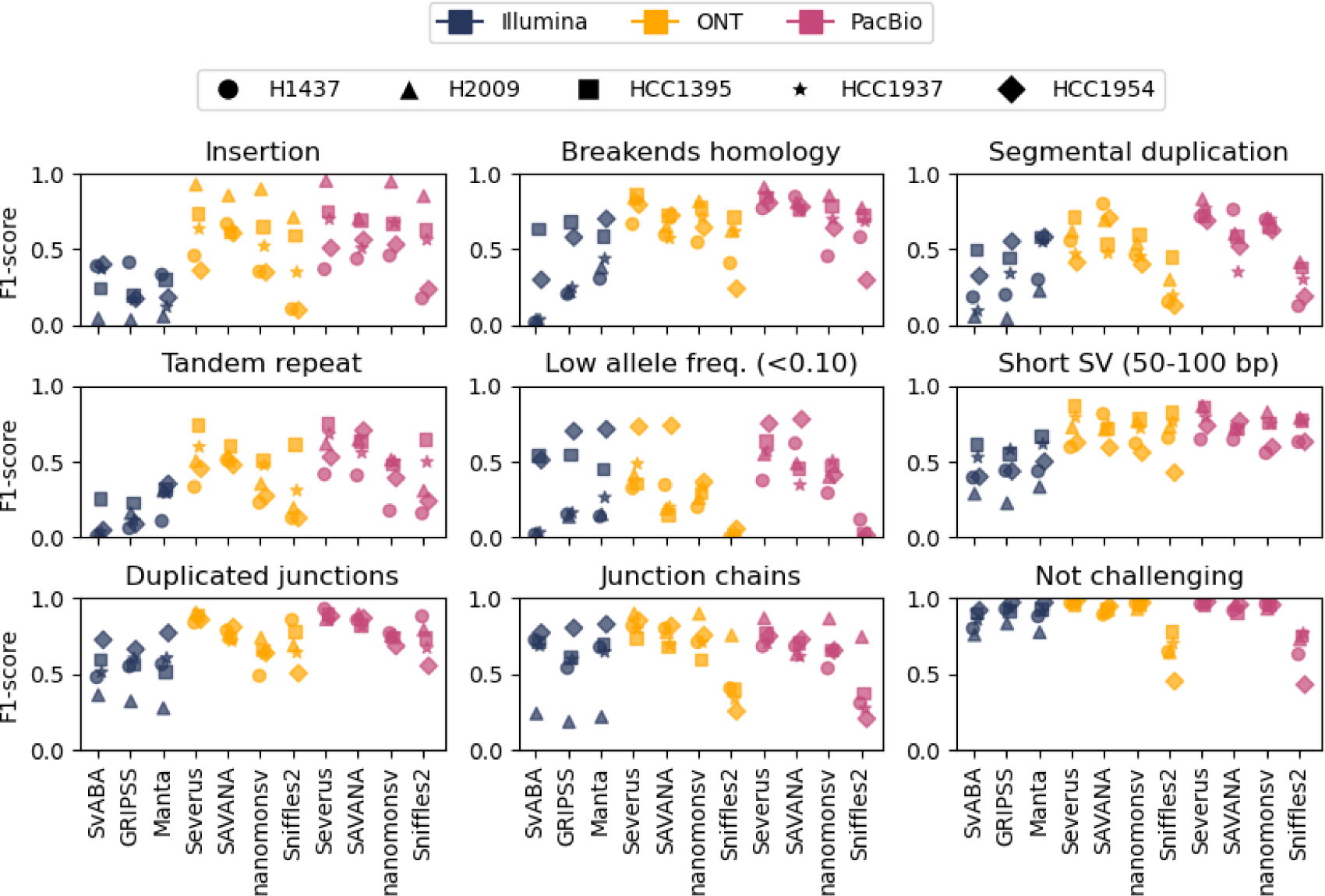
Stratification of error patterns of different sequencing technologies and algorithms. SV detection F1 scores are stratified by various challenging scenarios. Stratification and score computation were performed using Minda. Each cell line is shown with a different marker; colors show different sequencing technologies. Challenging categories may overlap. Repetitive genome regions are annotated using RepeatMasker tracks. Duplicates and rearrangement chains are defined for the union of all calls produced by all tools on a particular dataset. “Not challenging” reflects variants that do not belong to any challenging category.

As expected, insertions corresponded to a large portion of Illumina false-negative calls, and long-read methods had substantially better F1 scores associated with insertions. However, Sniffles2 and nanomonsv had an increased fraction of false-positive insertion errors, which could be explained by the disagreement in insertion calls between different long-read tools.

Next, we examined if junction breakpoint pairs were enriched with homologous repeats. We did not find such enrichment in true-positive SVs, consistent with the idea that homology-based mutational mechanisms may not be the primary source of somatic SVs in cancer (Choo et al. 2023). However, all Illumina methods had a substantial increase in false-positive calls with homologous repeat motifs. This suggests that repeat copies may cause read pair mismapping, leading to systematic false-positive calls.

As expected, all tools had more disagreements in calls with low VAF (<0.10). A substantial portion of these errors may be due to a failure to detect a true variant due to low read depth, especially in the case of PacBio (with 60x depth on average, compared to 100x for ONT and Illumina). Illumina methods also had an increase in false-negative calls for short SVs (50-100 bp) compared to the long-read methods.

In segmental duplication regions, SVaBA, nanomonsv, and Sniffles2 had an increased number of false-positive calls, which may be explained by mismapped short and long reads. All Illumina methods had a slight increase in false-negative errors in these regions, suggesting that short-read methods abstain from making calls in regions of low mappability. VNTR regions present a similar mappability challenge but at a higher magnitude, as many insertions occurred inside VNTR regions. On most datasets, Severus had one of the lowest error rates inside segmental duplications and VNTRs.

Finally, we considered chains of junctions as a challenging scenario. For example, a chain of breakpoint pairs (A -> B_1_) and (B_2_ -> C) could be erroneously called (A -> C) if the distance between B_1_ and B_2_ is small (Supplementary Figure 4). On most datasets, Illumina methods had an increased number of false-positive calls compared to the long-read methods. This may be explained by the difficulty of untangling clustered split-read connections. On the other hand, PacBio-based methods (but not ONT) had an increase in false negatives in SV chains. This reflects a tendency of the PacBio miniamp2 alignments to miss relatively short (100-500 bp) segments.

Importantly, most tools consistently had high F1 scores (>0.90) on SVs without apparent challenging patterns (Figure 4). This suggests that our stratification strategy represents the major difficulties in somatic SV calling for different technologies and tools.

### Benchmarking SV calling performance using downsampled data with variable tumor purity

In contrast to cell lines, tumor tissue samples often contain a fraction of normal cells, which represents an additional challenge for somatic variant callers. We sought to evaluate the performance of different methods and technologies using synthetically mixed samples with reduced tumor depth and purity. We used the set of confident SV calls from the HCC1954 genome as a baseline, as most methods performed well on this dataset. By mixing different proportions of reads sequenced from HCC1954 and HCC1954BL, we simulated 5 datasets with tumor purity varying from 20% to 100% for each sequencing technology.

As expected, all methods showed a reduction in F1 scores with decreasing tumor purity due to the reduced recall rate (Supplementary Figure 5; Supplementary Table 9). Within the PacBio and ONT categories, Severus consistently produced the highest F1 scores on all simulated datasets and improved over nanomonsv and SAVANA. SvABA showed F1 scores comparable to Severus, while GRIPSS and Manta had consistently lower performance. Severus had 62% recall on the ONT dataset with ∼16x haploid tumor depth and 45% recall on the PacBio dataset with ∼13x haploid tumor depth. Since the HCC1954 cell line is, on average, tetraploid, some somatic variants may be covered by only a few tumor reads, if any. This highlights the high sensitivity of Severus, while precision on most downsampled datasets remained >0.90.

### Patterns of complex rearrangements revealed by phased breakpoint graphs

We have now demonstrated that Severus produces accurate and comprehensive junction calls; however, cancer genomes are often characterized by complex mutational processes that produce many junctions simultaneously (Stephens et al. 2011). Severus enables the analysis of such complex events through the phased breakpoint graph framework, which aims to cluster junctions from the same complex events.

The majority of the SVs in cancer genomes are highly clustered yet consist mainly of simple SVs: deletions, insertions, tandem duplications, and reciprocal inversions (Y. Li et al. 2020). We define simple junctions as junctions that lead to simple SVs. Severus classifies complex (i.e., not simple) junctions into inversion-like and interchromosomal. Inversion-like events are further classified into reciprocal inversions with deletion/duplication, foldback inversions, BFB-like (multiple foldback inversions), and complex inversions (intermingled inversions). Interchromosomal junctions are categorized into reciprocal translocations, templated insertions, and translocation with inversion. Severus first filters junctions annotated as simple SVs and then constructs a breakpoint graph with the remaining complex junctions (Figure 5A). In the resulting graph, each connected component with 2+ junctions represents a complex SV (Methods, Supplementary Figure 6A).

**Figure 5.**
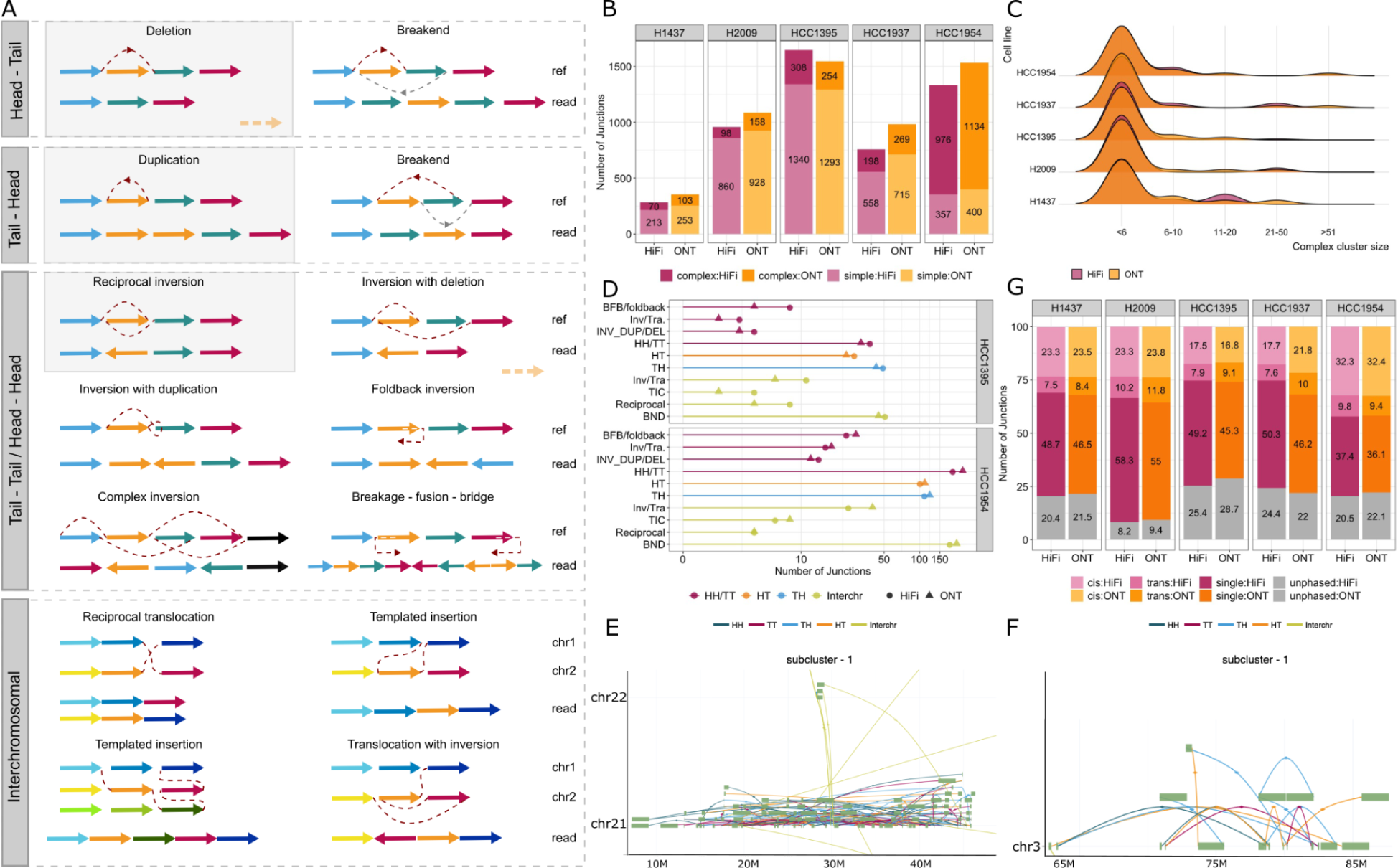
Overview of the complex SVs identified by Severus in a multi-technology cell line panel. (A) Examples of simple and complex SVs from each junction type; Head-to-tail (deletion-like), Tail-to-head (duplication-like), Head-to-head/Tail-to-tail (inversion-like), and Interchromosomal (translocation-like). Each colored arrow is a genomic segment, and the direction of the arrows represents the direction of the segment. Red dashed lines represent junctions. Simple SVs are indicated with a gray box and are not included in breakpoint graph construction. (B) Number of junctions in simple SVs and complex SVs in each cell line. (C) The distribution of complex cluster size in each cell line is colored by technology. (D) Number of junctions in each category involved in a complex SV. (E) A chromothripsis-like event in chr21 in HCC1954 and (F) in chr3 in HCC1395. (G) The distribution of junctions as unphased, a single phased junction within a phase block, and multiple junctions within a block; cis and trans refer to the same or different phases of two adjacent variants, respectively.

Using Severus, we observed characteristic numbers of complex SVs and determined SV cluster sizes among the cell lines. In all cell lines, the number of junctions in complex SVs was a smaller portion of the total number of junctions in that cell line (Figure 5B). HCC1954 was a clear exception, with the highest number of junctions in complex SVs, while HCC1395 and H2009 had the least. For all cell lines, the majority of complex SVs had 2-5 junctions. HCC1954 and HCC1937 had complex SVs with more than 20 junctions (Figure 5C).

Templated insertions and inversions with translocations were consistently found to be the most abundant SV types in all cell lines (Figure 5D, Supplementary Figure 6B, Supplementary Table 10). Consistently, the majority of the complex clusters had two to three chromosomes except for H1437, which had clusters within a single chromosome. Among the breast cancer cell lines, HCC1954, HCC1395, and HCC1937 exhibit similar mutational backgrounds and molecular classification, yet the complex SV clustering suggests different mechanisms might be involved in each cell line.

Amongst all cell lines, HCC1954, which has a highly rearranged karyotype, stood out as having the highest complex-to-simple ratio and an increased number of chromosomes within a single cluster. The largest cluster in HCC1954 has 337 junctions, 292 of them within or between chr5 and chr8. The translocation between chr5 and chr8 has been shown through FISH (Zhao et al. 2009), yet the complex nature of the rearrangement was unknown. Furthermore, the second largest cluster with 114 junctions in chr 21 was a chromothripsis-like rearrangement previously observed using Hi-C genome conformation capture (Akdemir et al. 2020) (Figure 5E).

HCC1395 had the smallest simple-to-complex ratio, where the majority of the events were non-reciprocal inversions and templated insertions and the largest cluster consisted of a chromothripsis-like event in chr3 causing rearrangements in *FOXP1* and *MAGI* (Figure 5F). We also observed chromoplexy in HCC1395 between chr2, 6, 8, 17, and 20 and in HCC1937 between chr1, 2, and 6 (Supplementary Figure 7). HCC1937 also had a higher number of templated insertion chain clusters compared to other cell lines.

In H2009, eleven clusters out of 22 exhibit a unique pattern as foldback inversion followed by a translocation (Supplementary Figure 7). We observed similar patterns in other cell lines yet to a lesser extent.

### Phased view of somatic SVs

Short-read SV clustering models cannot directly phase somatic SVs but rather operate under the assumption that clustered SVs appear in *cis*. This is, however, difficult to demonstrate directly without long-range phasing information. We therefore sought to confirm this model using the phased Severus calls. Within each phased block that contained more than one somatic SV, we counted the frequency of adjacent SVs either in *cis* or *trans* (Figure 5G). Overall, 25-40% of all SVs had at least one other SV within the phased block, consistent with the complex SV rates (Figure 5B). Among phased and clustered SVs, 75-85% of them were in the *cis* direction. A high proportion validates the idea of clustering junctions only within the same phase. Likewise, phasing provides the separation of relatively rare cases of unrelated SVs in close proximity but in different haplotypes.

### Long-read based clinical subtype stratification of pediatric leukemia cancers

We applied Severus to analyze SV in three clinical cases of pediatric cancer: two acute myeloid leukemia (AML) cases (CM1 and CM2) and one case of anaplastic large cell lymphoma (ALCL; CM3). All three cases had been subject to standard clinical testing workflows, which included karyotyping, custom FISH panels depending on disease type, SNP-based microarray for copy number variant detection, and DNA-based Illumina WGS (∼120x tumor / ∼40x normal) for small variant identification. CM3 was selected as a known positive SV case, CM1 as a known negative SV case, and CM2 as a case that was genetically undefined (no driver identified to date). The study investigators running Severus were initially blinded to the genetic results from the clinical workflows. Tumor and normal samples were sequenced using PacBio HiFi at 30x coverage in tumor and 15x coverage in normal. Normal samples were derived from a buccal swab for case CM1 and a remission peripheral blood sample for CM2; a normal sample was unavailable for CM3.

Severus identified 174 and 57 somatic SVs in samples CM1 and CM2, respectively; CM3 was processed in tumor-only mode (Figure 6A). In CM3, Severus identified a single reciprocal translocation, a clonal event between chr2 and chr5, leading to an *NPM1::ALK* fusion (Figure 6B). This finding was consistent with prior clinical karyotype analysis and FISH testing, which had identified a t(2;5)(p23;q35) involving the *NPM1* and *ALK* genes, a hallmark of ALCL. Severus did not find any complex clusters in sample CM1, which was consistent with prior clinical testing as this case of AML was known to be cytogenetically normal via karyotyping and also known to harbor biallelic mutations in *CEBPA* via Illumina sequencing. These analyses further support the high specificity of Severus.

**Figure 6.**
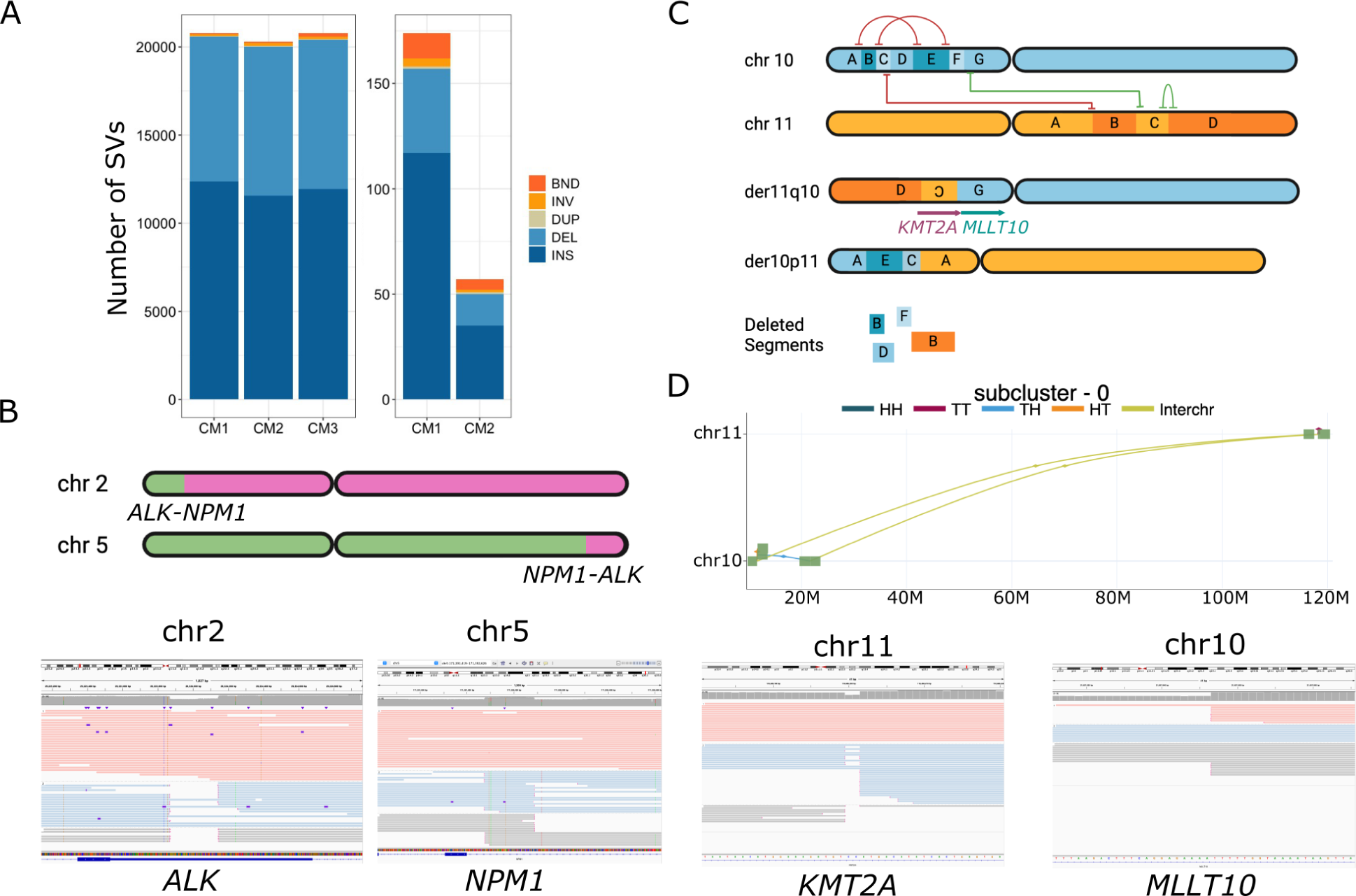
Severus identifies clinically relevant rearrangements in pediatric leukemia samples. (A) Number of germline (left) and somatic (right) SV calls in pediatric leukemia samples. (B) Reciprocal translocation event between chr 2 and chr 5 leading to an *NPM1::ALK* fusion in CM3. (C) Cryptic multi-breakpoint reciprocal translocation between chr10 and chr11 in CM2 leads to a *KMT2A*::*MLLT10* fusion. (D) Severus graph and IGV screenshots of translocation breakpoints in chr10 and 11. Reads are colored with the haplotype in IGV screenshots.

Finally, AML case CM2 was one that was ‘Not Otherwise Specified’ diagnostically (AML, NOS) and, despite prior clinical genetic testing (karyotype, FISH, microarray, and DNA-based sequencing), remained without an identified genetic driver. For this case, Severus identified a single complex cluster with five breakpoints involving chromosomes 10 and 11 that would be predicted to ultimately result in an in-frame *KMT2A*::*MLLT10* fusion (Figure 6C). The breakpoints detected by Severus were visually confirmed in IGV (Figure 6D). This fusion was not detected by prior karyotypic analysis nor FISH testing via a *KMT2A* break-apart probe, which is consistent with reports of this rearrangement being cytogenetically cryptic in some cases, as well as structurally complex, yielding false-negative FISH results (Peterson et al. 2019). Although not a part of the current analysis workflow, we ran Manta and GRIDSS. They identified 161 and 58 SVs between chr10 and chr11, whereas Severus reported only two (from the complex translocation event). GRIDSS also failed to identify the translocation leading to *KMT2A::MLLT10* fusion, which is possibly due to the close proximity between breakpoints in chr11. Overall, this analysis highlights the ability of long reads to aid in the resolution of genetically undefined pediatric leukemia cases while also confirming the high specificity of Severus.

## Discussion

In this manuscript, we introduced Severus - an algorithm for somatic SV calling and annotation using long reads. Severus takes advantage of better long-read mappability to call individual junctions and then builds a phased breakpoint graph to group junctions into complex SVs. The junction clustering problem is related to the problem of karyotyping, which has been previously studied using short-read methods (Hadi et al. 2020; Aganezov and Raphael 2020; Shale et al. 2022; Lee and Lee 2021). However, improved mappability and haplotype specificity of long reads resulted in less tangled SV graphs for Severus. It is possible that combining Severus with an orthogonal copy-number caller may further improve the karyotype reconstruction.

We evaluated Severus and other short- and long-read methods using synthetic and publicly available cell line benchmarks. In addition, we sequenced five commercially available tumor/normal cell line pairs using an array of sequencing technologies and generated consensus sets of benchmarking SV calls. Severus consistently had the highest F1 scores against this panel across all three PacBio, ONT, and Illumina methods. To perform these benchmarks, we developed a new comparison and merging tool called Minda with the improved support of complex somatic SVs.

To the best of our knowledge, our multi-technology cell line panel represents the most comprehensive benchmark for somatic SVs to date. Since we require the support of multiple tools and technologies for the variants in our cell line benchmarking set, it is possible that this strategy misses rare variants or variants with unusual patterns. This represents a tradeoff between the completeness of the benchmark and the practicality of its generation. We are publicly releasing all sequencing data and variant calls to allow manual curation of the disagreements between the different methods and technologies. Other research is underway to create high-quality benchmarks for somatic SV methods (Paulin et al. 2024). For example, the Genome in a Bottle consortium has recently released short- and long-read data of a pancreatic cancer cell line (https://www.nist.gov/programs-projects/cancer-genome-bottle).

We have also applied Severus to three pediatric leukemia/lymphoma cases. Severus was able to detect the diagnostic rearrangement in the known positive case and did not detect any diagnostic rearrangements in the known negative case. Finally, for the one genetically undefined case, Severus detected a cryptic diagnostic rearrangement (*KMT2A*::*MLLT10*) that was missed by FISH and karyotyping and was incomplete in Illumina SV calls. Identification of this fusion carries strong clinical significance as it confers a specific genetic subtype diagnosis for AML, is considered a high-risk fusion associated with poor prognosis, and alters the treatment approach by giving an indication for hematopoietic stem cell transplant and the addition of targeted therapy with Gemtuzumab (Pollard et al. 2021). This highlights the potential of the long-read whole genome sequencing approach for the diagnosis of complex cases driven by SVs.

Consistent with the previous reports (Choo et al. 2023), the performance of Illumina methods on somatic SVs was substantially better compared to the germline short-read benchmarks (Zook et al. 2020). This is primarily because a smaller proportion of somatic SVs are in repetitive regions. Nevertheless, long-read methods outperformed Illumina-based methods on most datasets. Our analysis explains most of the performance differences through multiple categories of challenging SV patterns. While some of the challenging cases present fundamental limitations for short reads (e.g., segmental duplications), other patterns (e.g., junction chains or short SVs) represent a potential opportunity for algorithmic improvements.

It is important to note that while we benchmarked individual SV calling tools, the ensemble approach is commonly used in large-scale studies (Y. Li et al. 2020). This approach can reduce the number of false-positive calls but, at the same time, increase the number of false-negative errors (Jeffares et al. 2017). Some SV calling pipelines can also incorporate additional genomic annotations (e.g., a panel of normals or blacklist). Evaluation of such database-enhanced strategies for SV detection is outside the scope of this work.

Because of the high heterogeneity of the SV landscape in cancer and a small sample size, our benchmarks may not represent the full complexity of the somatic SVs in different cancer types. However, the consistent high performance of Severus across all analyzed datasets is encouraging. Analysis of clinical tumor samples represents additional challenges, such as heterogeneity, tumor purity, and low biomass. Some of these difficulties are tangential to algorithmic developments, but nevertheless, testing on larger sets of clinical tumor samples is an essential next step. Fortunately, multiple ongoing initiatives (https://www.genomicsengland.co.uk/initiatives/cancer; https://www.bcgsc.ca/personalized-oncogenomics-program; https://www.childrensmercy.org/childrens-mercy-research-institute/studies-and-trials/genomic-answers-for-kids/) are currently aimed to generate long-read sequencing of various tumor types at scale.

## Acknowledgements

The work was supported in part by the Intramural Research Program of the NIH. This work utilized the computational resources of the NIH HPC Biowulf cluster. (http://hpc.nih.gov). ONT sequencing of the HCC1395 cell line was supported by the National Cancer Institute of the National Institutes of Health under Award Number U01CA253405. The content is solely the responsibility of the authors and does not necessarily represent the official views of the National Institutes of Health. The authors would like to thank the patients and families who donated their samples for this research. M.S.F. and E.G. would like to thank Braden’s Hope for Childhood Cancer, Elizabeth and Monte McDowell, the Black & Veatch Foundation, and Big Slick for their generous support. M.S.F., E.G., and L.L. would also like to thank Children’s Mercy Oncology Biorepository study personnel: Judy Vun, Amie Hatfield, and Robin Ryan; as well as Jason Seymour and Keiondra Sanders in the Children’s Mercy Research Institute (CMRI) Biorepository, for their assistance with sample collection and processing; and Maggie Gibson, Adam Walter, Laura Puckett in the CMRI Genomics Core for their assistance with sequencing. Y.L. is funded by the NCI-UMD Partnership Program. E.K.M. was supported by the State of Maryland. B.P. was supported by the National Human Genome Research Institute (NHGRI) under award numbers R01HG010485, U01HG013748, U24HG011853, U24HG010262, and U41HG010972, and from NIH award OT2OD033761.

## Competing interests

S.A. is an employee and stockholder of Oxford Nanopore Technologies. A.K., P.C., K.S., D.C., A.C. are employees of Google LLC and own Alphabet stock as part of the standard compensation package. E.G. served on advisory boards for Jazz Pharmaceuticals and Syndax Pharmaceuticals. M.S.F. is part of the speakers bureau for Bayer and PacBio. The remaining authors declare no competing interests.

## Methods

### Ethics statement

For the cell lines analysis, Institutional Review Board of National Institutes of Health considers patient-derived cell lines as non-human subjects, and no approval was required. There are, however, ethical considerations, as the cell lines were derived prior to establishing the research use consent mechanism, and no such consent was received. Commercially available cell lines used in this study are anonymized, and the risks of identifying original patients or their immediate family members are low. On the other hand, openly releasing this data will significantly benefit research into developing new methods for detecting somatic variants - a critical task in current and future precision cancer therapies. We concluded that the benefits outweigh the risks and followed the practices established by the NCI and NHGRI in the TCGA tumor cell line data release (https://www.cancer.gov/ccg/research/genome-sequencing/tcga/history/ethics-policies). For the three leukemia/lymphoma cases, patients were enrolled by Children’s Mercy Hospital (CMH) into its institutional Tumor Bank research study, which was approved by the CMH Institutional Review Board and included patient consent for the collection, processing, storage, and sequencing of patient samples.

### Cell culture of five tumor/normal cell line pairs

Cell lines were purchased from the American Type Culture Collection (ATCC). All cell lines were cultured according to ATCC handling guidelines at 37°C and 5% CO_2_. Cell counts were enumerated by Automated Cell Counter (BioRad, TC20), washed twice with Phosphate Buffered Saline (Gibco, 10010023), placed into 6 × 10^6^ cell aliquots, flash frozen in liquid nitrogen, and stored at -80°C. Cancer cell lines: HCC1954 (ATCC, CRL-2338), H2009 (ATCC, CRL-5911), H1437 (ATCC, CRL-5872), and HCC1395 (ATCC, CRL-2324). Normal cell lines: HCC1954 BL (ATCC, CRL-2339), BL2009 (ATCC, CRL-5961), HCC1937, BL1437 (ATCC, CRL-5958), and HCC1395 BL (ATCC, CRL-2325). Utilizing the Monarch® HMW DNA Extraction Kit for Tissue (New England Biolabs, T3060), high molecular weight (HMW) DNA was extracted from 6 × 10^6^ cells for all cell lines.

### ONT sequencing of five tumor/normal cell line pairs

HMW DNA was sheared to a target size of 50kb using the Megaruptor 3 and DNAFluid+ kit (Diagenode, E07020001). DNA was quantified using the dsDNA BR assay on a Qubit flourometer (ThermoFisher, Q33265), and size distribution was analyzed on Femto Pulse using the gDNA 165kb Analysis Kit (Agilent, FP-1002-0275). An SRE Kit (PacBio,102-208-300) was used to deplete DNA fragments <25kb in length. Libraries were created using an Ligation Sequencing Kit V14 (ONT, SQK-LSK114) following the standard kit protocol. Three library preps were conducted for cancer cell lines and one for normal cell lines. Cancer cell lines were sequenced on three flow cells, and normal cell lines were sequenced on one flow cell.

Libraries were eluted in 100µl of elution buffer. Sequencing was performed with version R10.4.1 PromethION flow cells (ONT, FLO-PRO114M) per manufacturer’s guidelines with the following alterations to maximize throughput; each library consisted of 25µL of DNA, 50µL of Load Beads, and 75µL of Sequencing Buffer to allow for four 150µl library loads per prep, and each flowcell was washed with the Flow Cell Wash Kit (ONT, EXP-WSH004) and reloaded every 24 hours, for a total runtime of 96 hours with four library loads.

### PacBio sequencing of five tumor/normal cell line pairs

Tumor and normal DNA was prepared for HiFi sequencing as previously described (Cohen et al. 2022) with the following modifications. Briefly, DNA was sheared to a target size of 14 kb using the Diagenode Megaruptor3, and SMRTbell libraries were prepared with the SMRTbell Express Template Prep Kit 3.0. Libraries were sequenced on Revio instrumentation (Pacific Biosciences, Menlo Park, CA) with 24 hr movies/SMRT cell. All samples were sequenced to a target depth of ∼60X coverage (2 SMRT cells per sample).

### Illumina sequencing of five tumor/normal cell line pairs

We used the TruSeq Nano DNA Prep from Illumina to prepare libraries. We took 200 ng of genomic DNA and fragmented it to a 400 bp insert size on a Covaris instrument which generates dsDNA fragments with 3’ or 5’ overhangs. The sheared DNA is blunt-ended and library size selection was performed using sample purification beads. A single ‘A’ nucleotide is added to the 3’ ends of the blunt fragments to prevent them from ligating to each other during the adapter ligation reaction. A corresponding single ‘T’ nucleotide on the 3’ end of the adapter provides a complementary overhang for ligating the adapter to the fragment. The indexed adapters are ligated to the ends of the DNA fragments and then PCR-amplified to enrich for fragments that have adapters on both ends. The final purified product is then quantitated by qPCR before cluster generation and paired-end sequencing (2 x150 bp) on the Illumina NovaSeq 6000 sequencer.

### ONT sequencing of HCC1395

The sample was fragmented to 20kb using Covaris g-TUBEs (COVARIS, 520079). DNA repair and end-prep was performed using the NEBNext Companion Module for Oxford Nanopore Technologies (NEB, E7180S) incubating at 20o C for 5 min followed by 5 min at 65o C. The sample was cleaned up using 1X Ampure XP beads (Beckman Coulter, A63881), washing twice on a magnetic rack with 80% ethanol. Sequencing adaptors and ligation buffer from the Oxford Nanopore Ligation Sequencing Kit (ONT, LSK1114) were ligated to DNA ends using Quick T4 DNA Ligase (NEB, E6056) for 45 min at room temp. The sample was cleaned up using 0.45X Ampure XP beads (Beckman Coulter, A63881), washing twice on a magnetic rack with the long-fragment buffer (ONT, LSK114) before eluting in 32uL of elution buffer (ONT, LSK114). Sequencing libraries were prepared by adding the following to the eluate: 100uL sequencing buffer (ONT, LSK114), 68uL loading solution (ONT, LSK114), and 0.5uL sequencing tether (ONT, LSK114). Samples were run on a PromethION (ver R10.4.1) flow cell using the PromethION sequencer. Sequencing runs were operated using the MinKNOW software (ver 22.12.5).

### ONT and Illumina sequencing of COLO829/COLO829BL

For ONT sequencing, DNA extraction was performed using the QIAGEN Puregene Cell Kit as recommended by Oxford Nanopore Technologies starting with 5 × 10⁶ COLO829 and COLO829 BL cells. Size-selection was performed on the COLO829 BL DNA following the Size selection of HMW DNA by semi-selective DNA precipitation protocol. Libraries were created using the Ligation Sequencing Kit V10 (ONT, SQK-LSK110) following the standard kit protocol. Libraries were sequenced in 24µl Elution Buffer (EB), 75 uL Sequencing Buffer (SBII) and 51 uL Loading Beads (LBII) on R9.4.1 PromethION flow cells as per manufacturer’s guidelines. To maximize throughput each flow cell was washed with the Flow Cell Wash Kit (ONT, EXP-WSH004) and reloaded every 24 hours, for a total runtime of 72 hours with three library loads.

For Illumina, cell lines gDNA was sequenced via Source Bioscience sequencing service provider. PRC-free library preparation with KAPA HyperPlus Kit was performed. Prepared libraries were sequenced on Illumina NovaSeq 6000 sequencer. Raw fastq files were then received and utilized in the subsequent analysis.

### Pacbio HiFi sequencing of pediatric leukemia samples

Samples libraries were prepared and sequenced as previously described (Cohen et al. 2022). Libraries were sequenced on the Sequel IIe System with 30 hour movies/SMRT cell to a target depth of coverage of ∼24x (3 SMRT cells per sample, tumor) or ∼16x (2 SMRT cells per sample, normal).

### Severus algorithm and rearrangement model

Below, we provide a detailed description of the Severus algorithm. It consists of two major parts: (i) junction signatures extraction and (ii) SV discovery using a breakpoint graph.

A junction brings together two non-adjacent reference locations called breakpoints (at leads BP_DISTANCE apart or on different chromosomes). Each breakpoint is directional, either *head* or *tail* (corresponding to 3’ and 5’ directions, respectively). Thus, a junction is defined by an ordered pair {*(start_position, start_direction), (end_position, end_direction)}*, where *start_position* < *end_position*. For example, a deletion at position *P* of size *L* corresponds to: *(P, head):(P + L, tail).* Unmapped insertions are an exception, as they are defined by a single coordinate. A junction may be copy-neutral (translocation) or result in a decrease (deletion) or increase (duplication) in the number of copies.

### Alignment, phasing, and junction signatures detection

A key advantage of long-read sequencing is the ability to phase variants into longer haplotypes without the need for any additional data. Heterozygous germline variants are typically more frequent by orders of magnitude than somatic variants; we thus focus on separating two germline haplotypes in both tumor and normal samples. If the normal sample is available, we call and phase SNPs in the normal sample using DeepVariant and Margin (Shafin et al. 2021). We then assign haplotypes for aligned reads in both normal and tumor samples using Whatshap (Martin et al. 2016). Some reads may remain unphased if they do not span any heterozygous variants. In the absence of a normal sample, phasing of tumor alignment data can be performed instead.

Similar to other long-read SV calling tools, Severus then extracts junction signatures from (i) reads with split alignments or (ii) reads with long indels inside a single alignment. In some cases, a junction may be represented by reads with both (i) and (ii) signature types. Below, we explain how these signatures are preprocessed and used to build the breakpoint graph.

In some cases, primary and supplementary alignments may overlap in read coordinates. This may happen if both breakpoints are flanked by a repeat - a common signature of reciprocal inversions. If two read alignments overlap in both read and reference coordinates, one of the alignments is trimmed to remove the overlap.

### Synchronizing indel signatures inside VNTR regions

Recent studies highlighted that variation inside variable number tandem repeats (VNTRs) is one of the major sources of error in SV analysis (Kolmogorov et al., 2023). This is because the alignment inside the VNTR regions is often ambiguous and may have multiple near-optimal solutions; as a result, alignments become sensitive to read errors. This is problematic because the same SV may be detected differently in tumor and normal samples, resulting in a false positive somatic call.

Most SVs inside VNTRs are indels (Chaisson et al. 2019). For each read, Severus clusters indels inside the same VNTR region and represents it as a single event of the total corresponding length. In addition, an insertion may be represented as a duplication (and vice versa). In this case, it corresponds to a split alignment with two parts that overlap in reference coordinates. Severus converts the split-read signature into an insertion signature so that it can be matched with other reads.

Severus also handles interspersed indels outside the annotated VNTRs. We combine (i) deletions separated by a small (<500 bp) segment and (ii) insertions separated by a segment shorter than the cumulative length and represent it as a single signature with the cumulative length.

To enable VNTR signatures clustering, a user needs to provide a bed file with VNTR annotations. It could be performed de novo using the “findTandemRepeats” script from the pbsv package (https://github.com/PacificBiosciences/pbsv/tree/master/annotations). Annotations for the most popular reference genomes are provided in the repository.

### Detection of mismapped reads and collapsed duplications

Although long-read sequencing dramatically improved read mappability throughout the genome compared to short reads, very long repeats (such as segmental duplications) still present a mapping challenge. Mismapped reads from wrong repeat copies may artificially inflate (or deplete) the coverage of the region, and these reads may contain junction signatures, resulting in false-positive calls.

Mismapped repeats are more prevalent in non-human genome alignments (e.g., against a mouse reference genome) since non-human reference genomes typically have lower assembly quality and a higher rate of collapsed duplications.

Since the copies of evolutionary divergent repeats are not identical, a common signature of mismapped reads is an increased sequence divergence, as compared to the correctly mapped reads. In Severus, we are using a read substitution rate above the baseline as a filtering criterion. The threshold depends on the error rate of the input reads and is computed as 0.95 quantiles of all reads, providing flexibility to work with technologies with different error profiles (e.g., ONT R9, ONT R10, PacBio HiFi). Since some regions can be hypermutated by nature (e.g., HLA region), we identify regions containing reads with lower and higher than the substitution rate threshold (at least 10% each). All reads within those regions and substitution rates above the threshold are considered mismapped and called failed.

### Selecting reliable reads

Similarly to other structural variant callers, we apply additional filters to individual reads to further control for possible read mismapping. For each read, Severus requires (i) minimum alignment length (N90 by default), (ii) minimum aligned ratio (0.5), and (iii) minimum mapping quality (MAPQ; 10 by default). Reads that fail at least one of these tests are called failed. Severus also outputs the number of ‘PASS’ and ‘FAIL’ reads as well as the total length of each category.

Filtering individual reads may not be sufficient and misleading, as a few reads with borderline quality values may still support a junction signature, even if the majority of reads have low quality and uncertain alignments. Therefore, instead of removing failed reads, we use all reads to generate junction signatures. Afterward, we compute the rate of failed reads that support a signature. If the rate is higher than the 0.5 threshold, the signature is marked as unreliable and is not used to generate the final output.

### Breakpoint graph framework

The breakpoint graph is a popular framework for studying genomic rearrangements in comparative genomics (Alekseyev and Pevzner 2009) and cancer genomics (Malhotra et al. 2013). In a popular definition, nodes in the breakpoint graph correspond to the SV breakpoints. The nodes are connected by two types of edges: adjacency (corresponding to SV junctions) and genomic (connecting adjacent breakpoints in the reference). In other works, a genomic edge corresponds to a maximum region of the reference genome that is free of rearrangements.

In this work, we use an equivalent definition of a genomic segment, that corresponds to a pair of breakpoints connected by a genomic edge (similarly to sequence graphs in a GFA format). Two segment extremities (start and end) correspond to the original breakpoints. Therefore, a breakpoint graph is a collection of genomic segments that are connected by adjacency edges (Figure 1).

### Defining adjacency edges

Given a set of junction signatures extracted from alignments, Severus builds a breakpoint graph as follows. First, junctions are clustered based on their start breakpoint positions and direction (within BP_DISTANCE = 50 bp by default). Within this initial cluster, we perform a second round of clustering based on the end breakpoint. Each secondary cluster is accepted if (i) it is supported by more than BP_MIN_SUPP reads (3 by default) and (ii) the rate of failing supporting reads is lower than BP_MAX_FAIL (0.5 by default). For each accepted cluster, we create two nodes in the breakpoint graph (for start and end breakpoints) and connect the nodes with an *adjacency* edge (which holds additional information, such as sample ID and number of supporting reads).

Therefore, a junction is defined by a pair of two breakpoints connected by at least one adjacency edge, and in the case of multiple simple analyses, multiple parallel adjacency edges may be created. If a junction is supported by a matching normal sample with a frequency higher than MAX_TIN (0.01 by default), the junction is labeled germline and otherwise somatic.

### Defining genomic segments

The goal of the next stage is to determine the arrangement of breakpoints in the derived chromosome configuration (and specific for each germline haplotype). In a standard breakpoint graph definition, genomic edges are created for all pairs of breakpoints with corresponding directions that are adjacent in the reference coordinates; however, it assumes that all junctions are copy-neutral. In addition, this ignores the connectivity information from long reads that span multiple junctions.

In Severus, we add genomic edges to the graph in two stages as follows. In the first stage, for each read that spans at least two junctions - defined by 4 breakpoints {(A, B), (C, D)} - we add a genomic edge between the intermediate breakpoints (B and C). After all reads are processed, we consider breakpoints that do not yet have an incident genomic edge. For each such breakpoint, we search for a mate breakpoint with (i) consistent direction, (ii) within MAX_GENOMIC_DIST (2 Mb by default) and (iii) consistent coverage between breakpoints.

Note that the first iteration of the genomic edge algorithm is important, as read-based genomic connections always have priority over reference-based connections. The MAX_GENOMIC_DIST parameter ensures the separation of unrelated distant SV clusters. In case a region is unphased, we do not check phase consistency for genomic edges. In case a genomic edge spans a phase block boundary, we add a special type of adjacency edge that indicates that.

Genomic edges and their incident breakpoints are then converted into genomic segments. Note that genomic segments may overlap in their coordinates because of (i) SVs in close proximity but on different haplotypes (ii) resolved amplifications (iii) heterogeneity. Severus constructs two separate breakpoint graphs: with only somatic SVs and with all SVs.

### Additional clustering and annotation of complex SVs

For the complex SV clustering, we first filter simple SVs that are less likely to be part of a complex event: small deletions (<10 kb), insertions, small tandem duplications (<10 kb), and reciprocal inversions (often referred to as balanced inversions; Figure 5A). Severus then constructs a breakpoint graph with the remaining SVs.

The connected components of the constructed breakpoint graph already represent clusters of SVs, for example, chains of deletions or translocations. Therefore, it automatically groups breakpoints of complex SV events. However, in some cases the graph-based clustering fails to capture more complex SV patterns, either because of overlapping SVs or because of complex SV definitions. For example, chromoplexy is defined by multiple interchromosomal translocations, but the derived chromosome structures may be clustered into separate connected components. Another example is a Breakage-Fusion-Bridge amplification, which is flanked by foldback inversions but may contain more SVs inside the amplified regions.

To account for this, Severus implements additional clustering rules for several complex SV types. We cluster (i) inversions (head-to-head and tail-to-tail adjacencies) to identify reciprocal (canonical, with deletion, with duplication), non-reciprocal, foldback, and complex (with multiple inversions in a single path) inversions, which are often key players in complex events, (ii) breakpoints to identify reciprocal translocations, tandem duplications, and templated insertions. A subgraph with more than two SVs is defined as a complex SV cluster and assigned a cluster ID.

### Generating output

Severus generates both graph and VCF outputs. One or more adjacency edges correspond to a VCF entry (for example, one adjacency for deletion and two for reciprocal inversion). SVs identified as part of a complex event contain a cluster ID corresponding to the SV signature grouping described above. Severus also outputs the list of breakpoints with additional information (e.g., sample genotypes and supporting reads) in a CSV format. Severus provides the graphs as (i) a graphviz network and (ii) an interactive Plotly plot.

### Minda algorithm

Recently, a number of tools for SV benchmarking and merging have been developed, such as truvari, SVanalyzer, SURVIVOR, and Jasmine. However, these tools were primarily designed for the processing of germline indel variants and do not support other types of SVs. Another challenge is that different SV calling tools represent the same SVs in VCF differently.

Thus, for our evaluations, we developed a new tool called Minda that unifies an input VCF by decomposing every SV into start and end records. Minda clusters two SV records together, if the corresponding starts and ends are within the TOLERANCE threshold (500 bp by default). Minda then estimates the recall and precision against the defined ground truth set of SVs. In the ensemble mode, Minda merges multiple sets of SVs, and then each SV cluster is classified as confident if it is supported by the given number of technologies and callsets. Recall and precision are then calculated against this confident SV set.

Minda can also be used for multiway comparison of multiple sets of SV calls and SV merging, as other tools with merging functionality - such as SURVIVOR (Jeffares et al. 2017) or Jasmin (Kirsche et al. 2023) - were tailored for germline variation analysis.

To ensure consistency, we compared Minda against truvari on the HG002 GIAB SV benchmark using a 500 bp tolerance and 50 bp minimum SV length. Both tools had comparable recall and precision values (Supplementary Figure 1; Supplementary Table 4). In contrast, on the CHM1/CHM13 (500 bp tolerance and 50 bp minimum SV length) and COLO829 (500 bp tolerance and 30 bp minimum SV length) benchmark sets, truvari output lower scores for some tools (Supplementary Table 3). Some vcf files required preprocessing to run truvari without errors. The vcf produced by Valle-Inclan et al., (2022) and SAVANA vcf required INFO fields to be appended with SV end coordinates. Though truvari ran without producing an error on SvABA vcf files, no calls were identified.

### Basecalling, alignment and phasing

For ONT samples, Guppy (v6.3.9) was used for basecalling and followed by alignment to hg38 genome using minimap2 v2.24 (H. Li 2021) with ‘*-k 17 -y –eqx*’ parameters. Normal samples were subjected to the Pepper-DeepVariant-Margin v0.8 (Shafin et al. 2021) pipeline for phasing. The subsequent phased vcf used to haplotag both normal and tumor samples using Whatshap v1.7 (Martin et al. 2016). HiFi reads were aligned using minimap2 with the same parameters to hg38 genome. The phased vcf generated with ONT data were used to haplotagged normal and tumor samples. Illumina reads were aligned using bwa-mem2 v2.2.1 (H. Li and Durbin 2009) with default parameters to hg38 genome.

### Running SV detection tools and benchmarking

Severus (version 0.1.2), SAVANA (version 1.0.3), Manta (version 1.6.0), SvABA (version 1.2.0), and GRIDSS (version2.13.2) followed by GRIPSS (version 2.2) were run with tumor and normal bam pairs using default settings. For Severus, inputs also included a phased vcf described above and tandem repeat annotations file was downloaded from https://github.com/PacificBiosciences/pbsv. “nanomonsv parse” (version 0.6.0) was run with default parameters, and “nanomonsv get*”* was run with *--use_racon* and *--single_bnd* options. For Sniffles (version 2.0.7), default settings were used for the normal alignment, while the *--non-germline* option was used for the tumor alignment. The normal and tumor “snf” output files were merged into a single vcf and parsed for calls only found in the tumor sample. Both tumor and normal Sniffles runs also used the tandem repeats annotation file as an input.

As the GIAB HG002 benchmark was originally designed for the grch37 human reference, we lifted over the benchmarking Tier1 confidence intervals to grch38 and created a ground truth SV set from a curated HG002 assembly (Liao et al. 2023) using hapdiff (Kolmogorov et al. 2023).

For the complex SV analysis Severus (v1.0) was run with ‘--resolve-overlap’ and ‘--single-bp’ parameters for improved clustering.

### Code availability

Severus is available at: https://github.com/KolmogorovLab/Severus. Minda is available at: https://github.com/KolmogorovLab/minda.

### Data availability

Cell line sequencing produced in this study is openly available at NCBI SRA BioProject PRJNA1086849. Accession codes for each individual sample are given in Supplementary Table 2. Sequencing of the clinical samples is under controlled access and will be available through dbGaP. Accession codes and links to the publicly available datasets used in this work are available in Supplementary Table 2. The outputs of all tools and Minda evaluations are available at: https://zenodo.org/doi/10.5281/zenodo.10856827.

## Supplementary Figures

**Supplementary Figure 1.**
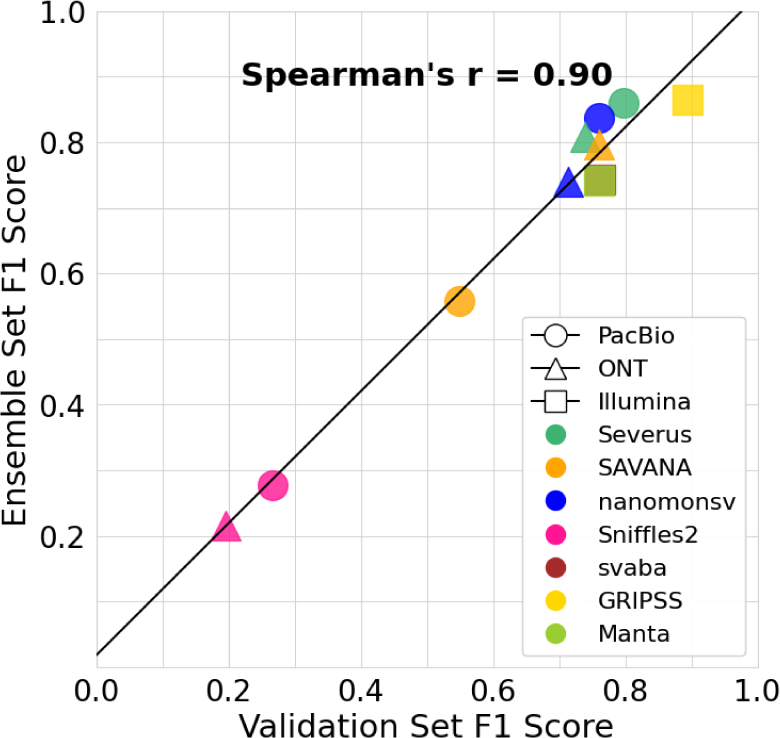
Consistency of the COLO829 Valle-Inclan benchmark and Minda ensemble benchmark.

**Supplementary Figure 2.**
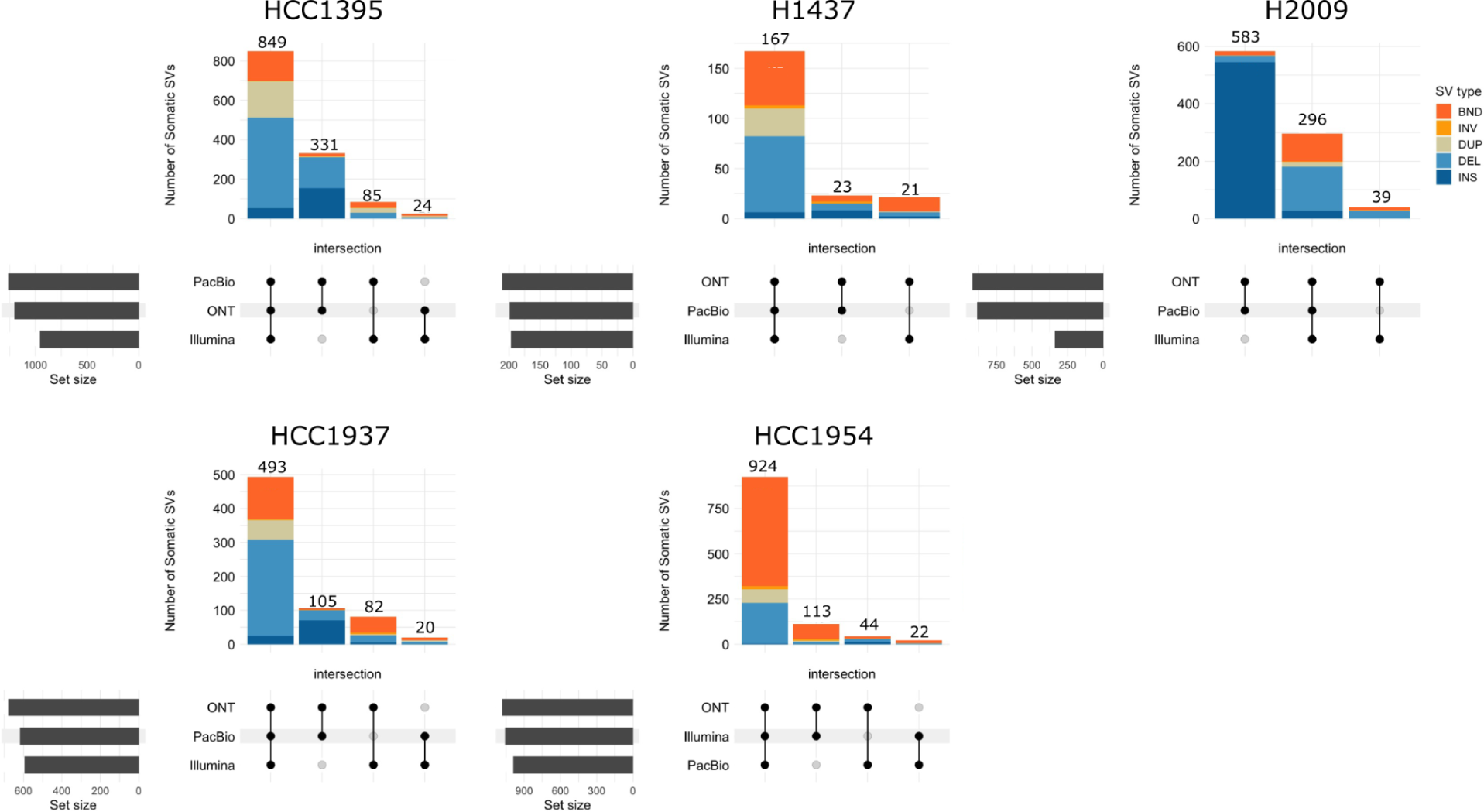
Similarities between SV calls produced by the different technologies. DEL = Deletion, BND = breakend junction, DUP = duplication, INV = inversion, INS = insertion.

**Supplementary Figure 3.**
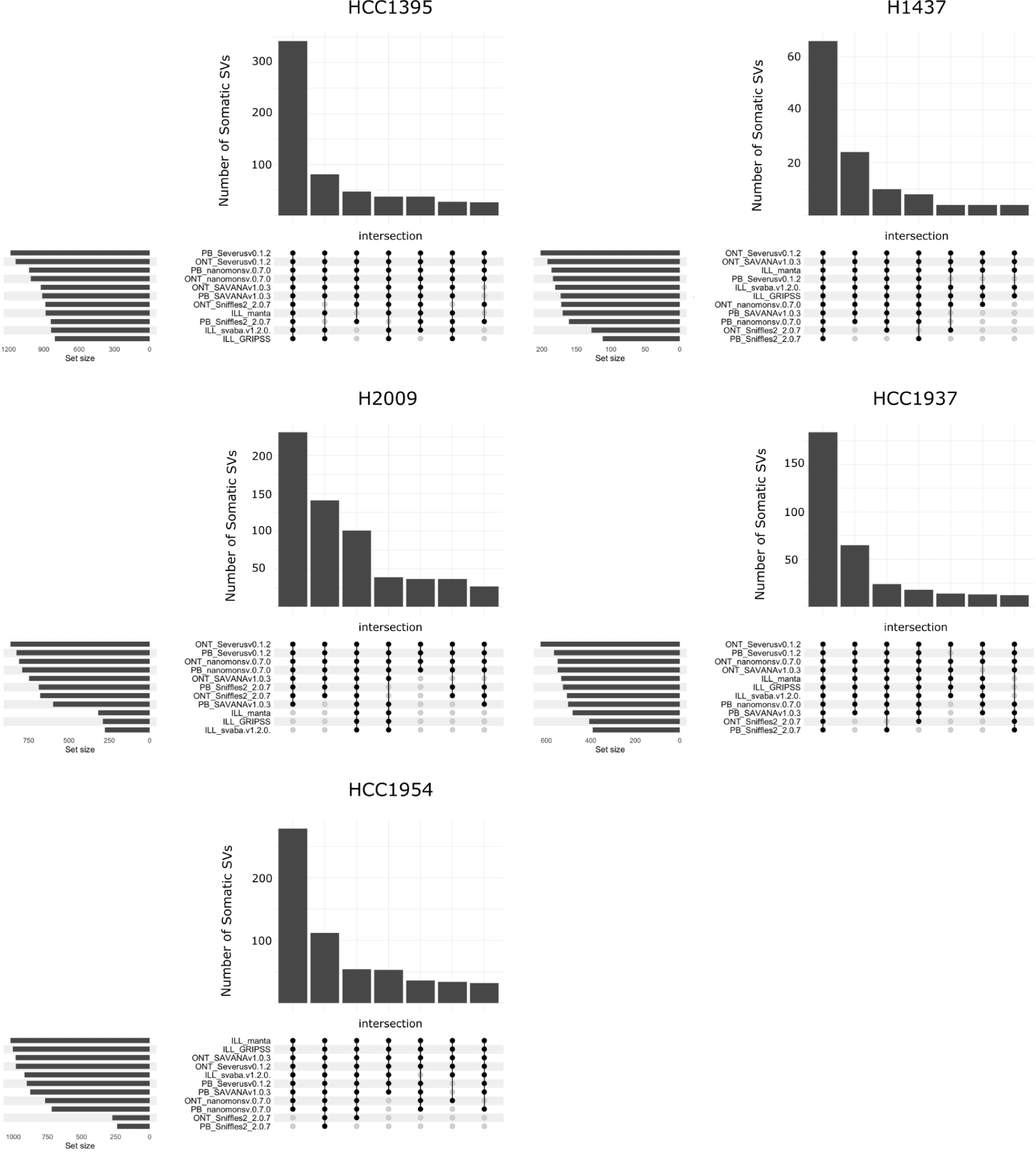
Similarities between SV calls produced by the different tools.

**Supplementary Figure 4.**
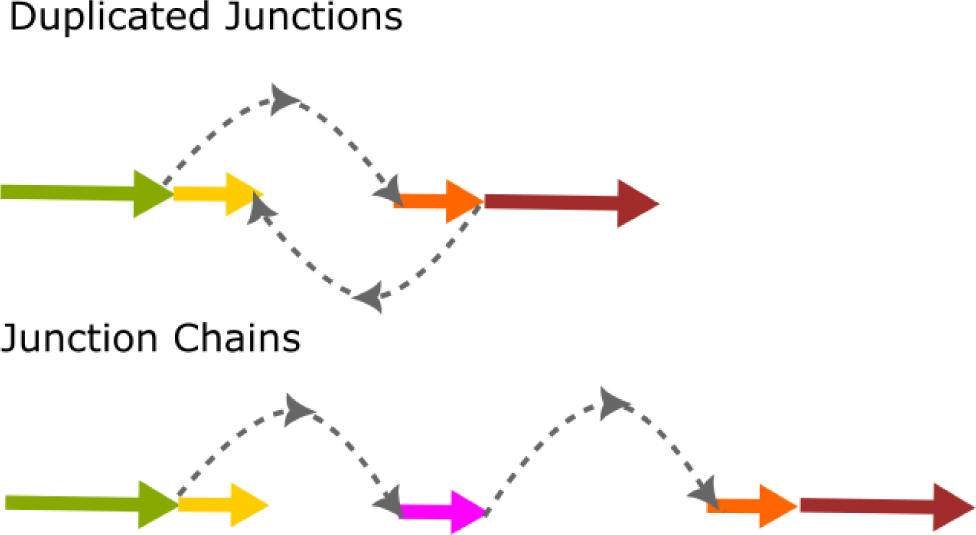
An example of a duplicated junction and a junction chain.

**Supplementary Figure 5.**
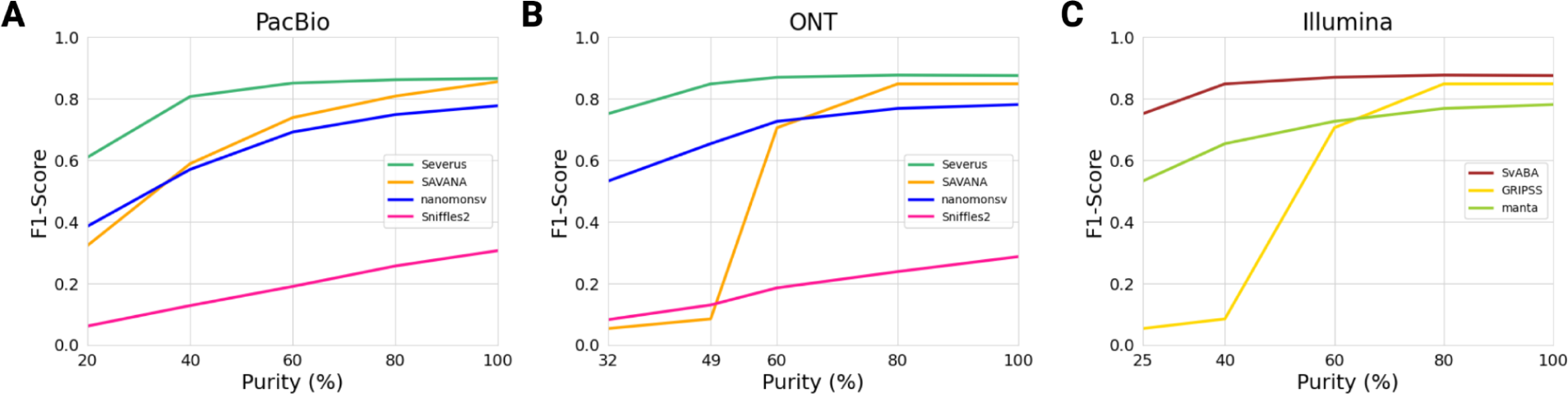
Performance of SV calling tools with variable levels of tumor purity and normal contamination.

**Supplementary Figure 6.**
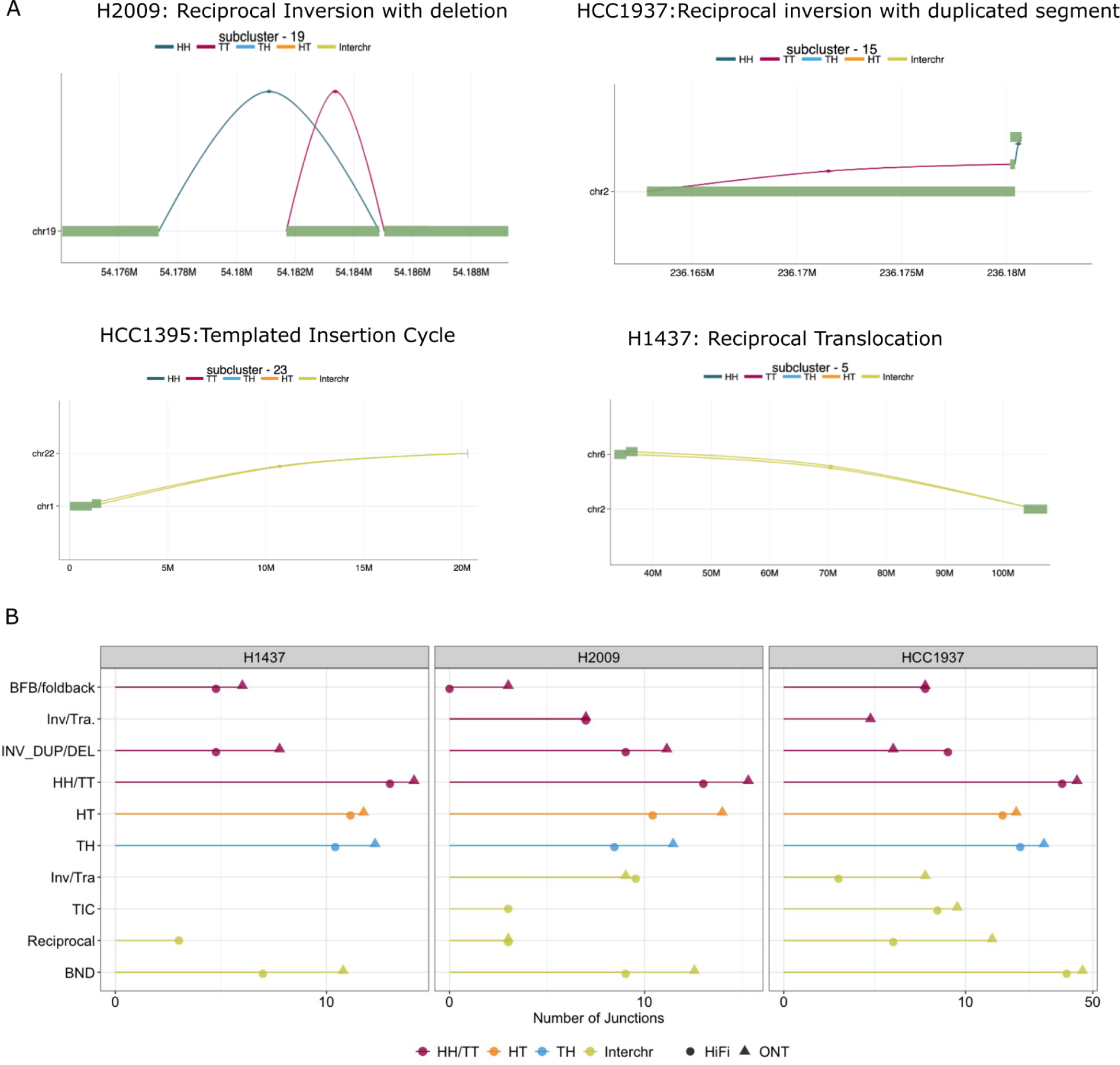
A. Example of graphs for detailed type annotations provided by Severus. B. Number of junctions in each subcategory involved in a complex SV.

**Supplementary Figure 7.**
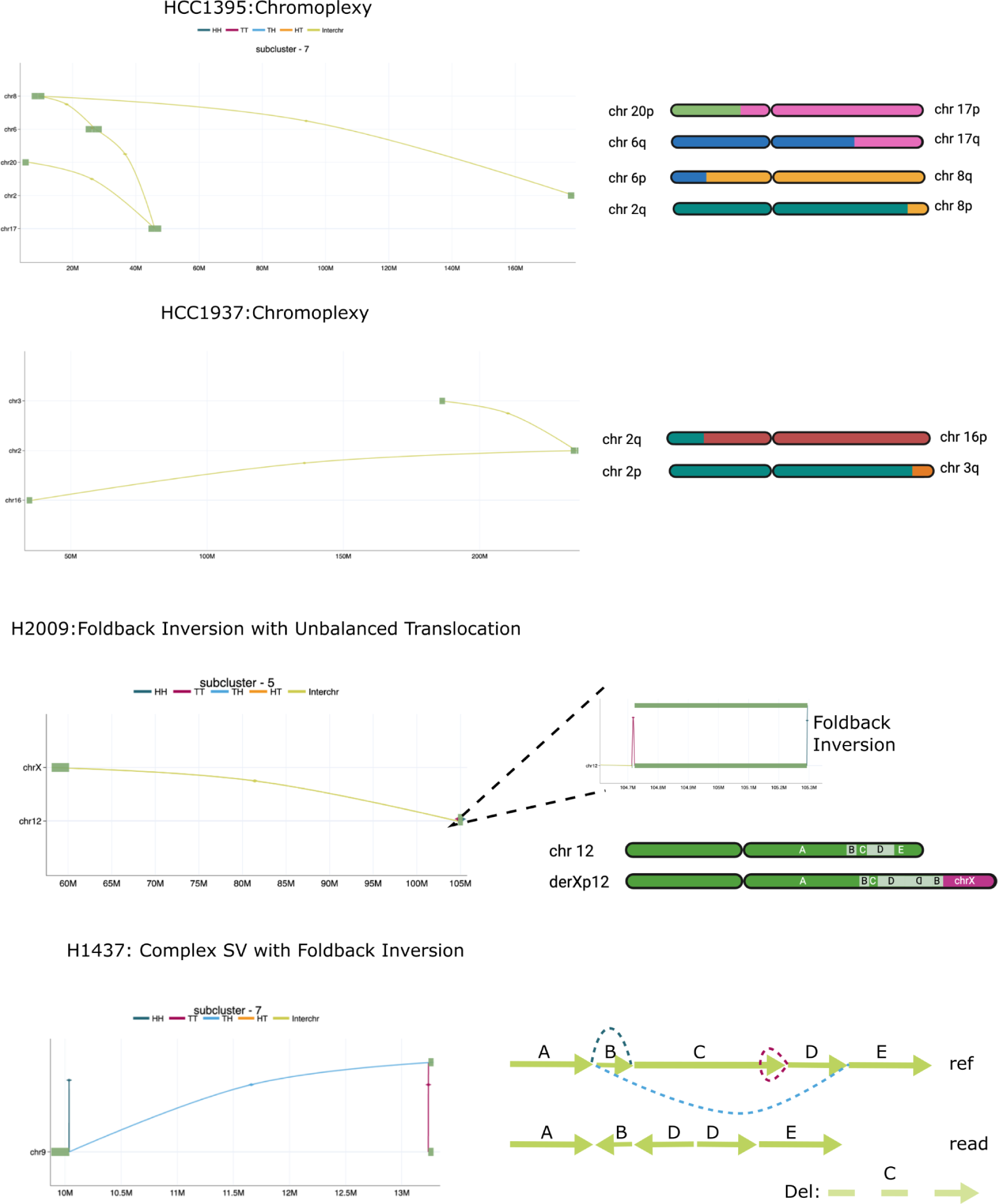
Additional examples of complex SVs discovered by Severus.

## Supplementary Tables List

**Supplementary Table 1:** Detailed statistics for the HG002 and CHM1/CHM13 benchmarks

**Supplementary Table 2:** Sequencing information and accession codes

**Supplementary Table 3:** Detailed statistics for COLO829 benchmarking results

**Supplementary Table 4:** Comparison between the COLO829 truth set and the ensemble approach

**Supplementary Table 5.** Sensitivity of the ensemble call set variation in Minda parameters on the HCC1395 dataset.

**Supplementary Table 6**. Detailed statistics for the cell line panel ensemble benchmark.

**Supplementary Table 7.** Analysis of Severus results on downsampled data from HCC1954 and H2009 samples.

**Supplementary Table 8:** Error patterns in different callers and technologies

**Supplementary Table 9:** Benchmarking using variable tumor purity.

**Supplementary Table 10:** Summary of complex events in cell lines

